# Discovering prescription patterns in pediatric acute-onset neuropsychiatric syndrome patients

**DOI:** 10.1101/19004440

**Authors:** Arturo Lopez Pineda, Armin Pourshafeie, Alexander Ioannidis, Collin McCloskey Leibold, Avis L. Chan, Carlos D. Bustamante, Jennifer Frankovich, Genevieve L. Wojcik

**Affiliations:** Department of Biomedical Data Science, Stanford University, CA, USA; Department of Physics, Stanford University, CA, USA; Department of Pediatrics, Division of Allergy, Immunology, and Rheumatology, Stanford University, CA, USA; Department of Medicine, University of Massachusetts Medical School. Worcester, MA, USA; Department of Genetics, Stanford University, CA, USA; Chan Zuckerberg Biohub, San Francisco, CA, USA; Department of Epidemiology. Bloomberg School of Public Health. Johns Hopkins University, Baltimore, MD, USA

**Keywords:** Cluster Analysis, Patient similarity, Longitudinal Studies, Polypharmacy, Pediatric acute-onset neuropsychiatric syndrome

## Abstract

**Objective:** Pediatric acute-onset neuropsychiatric syndrome (PANS) is a complex neuropsychiatric syndrome characterized by an abrupt onset of obsessive-compulsive symptoms and/or severe eating restrictions, along with at least two concomitant debilitating cognitive, behavioral, or neurological symptoms. A wide range of pharmacological interventions along with behavioral and environmental modifications, and psychotherapies have been adopted to treat symptoms and underlying etiologies. Our goal was to develop a data-driven approach to identify treatment patterns in this cohort.

**Materials and Methods:** In this cohort study, we extracted medical prescription histories from electronic health records. We developed a modified dynamic programming approach to perform global alignment of those medication histories. Our approach is unique since it considers time gaps in prescription patterns as part of the similarity strategy.

**Results:** This study included 43 consecutive new-onset pre-pubertal patients who had at least 3 clinic visits. Our algorithm identified six clusters with distinct medication usage history which may represent clinician’s practice of treating PANS of different severities and etiologies i.e., two most severe groups requiring high dose intravenous steroids; two arthritic or inflammatory groups requiring prolonged nonsteroidal anti-inflammatory drug (NSAID); and two mild relapsing/remitting group treated with a short course of NSAID. The psychometric scores as outcomes in each cluster generally improved within the first two years.

**Discussion and conclusion:** Our algorithm shows potential to improve our knowledge of treatment patterns in the PANS cohort, while helping clinicians understand how patients respond to a combination of drugs.

## 1. Introduction

In cohorts of patients with multiple medication usage –polypharmacy– it is important to understand if there are groups of patients that share similar patterns of usage and what are the differences between these groups of patients. Polypharmacy is common in older patients with multimorbidity [1]; and it is associated with adverse outcomes including mortality and adverse drug reactions [2], increased length of stay in hospital and readmission to hospital soon after discharge [3]. Polypharmacy can also occur in children and adolescent patients with psychiatric diseases [4], and other non-elder adults with complex chronic syndromes such as lupus [5], human immunodeficiency virus (HIV) infection [6], ischemic and respiratory diseases [7], or cancer [8]. These diseases are often multifactorial, with physicians treating related, but separate, symptoms and pathologies. One of these syndromes is the pediatric acute-onset neuropsychiatric syndrome (PANS).

The PANS clinical presentation [9] is characterized by abrupt-onset of obsessive-compulsive disorder and/or food restriction, along with at least two other severe neuropsychiatric symptoms from the following categories: anxiety; mood lability or depression; irritability, oppositionality, or rage; behavioral regression; deterioration in school performance/cognitive difficulties; sensory or motor abnormalities; and somatic symptoms like sleep disturbances or enuresis. Patients typically experience a relapsing-remitting course in which disease flares are interspersed with remissions [10]. In some cases, the disease course is chronic, when the patient’s neuropsychiatric status does not return to baseline.

Current evidence suggests that PANS has an inflammatory or autoimmune etiology that is associated with an infection [11-15]. A recent MRI study in children with PANS showed an increased median diffusivity in multiple brain structures including basal ganglia, thalamus and amygdala compared to controls, suggesting neuroinflammation in these regions. Multidisciplinary clinics are well-positioned to care for patients with PANS [16]. Although treatment protocols are lacking, interim guidelines suggest using antibiotics to treat or prevent infections, immunomodulatory therapies to manage inflammation, and psychiatric medications supplemented with cognitive behavioral therapy to treat PANS [17]. The heterogeneity and complexity of PANS presentation, clinical course, treatment, insurance status, and irregular follow-up make it difficult to compare treatment courses across patients and patient-groups, necessitating a novel method to cluster patients while considering temporality.

In precision medicine, patient similarity is an emerging concept that aims to help discover groups of patients (clusters) that share similar characteristics estimating a numerical distance between components of patient data [18]; and ultimately, to use those clusters in predictive modeling tasks [19]. The data features commonly used include demographics and population characteristics [20], genetics [21], prescriptions and lab tests [22, 23], medical billing codes [24], and even clinical narratives processed with natural language tools [25]. Meanwhile, commonly used algorithms for this task employ distance-based functions that consider geometrical space (e.g. Euclidean, Manhattan, etc.) [22, 26]; machine learning models that employ representations of patients in the multivariate space (e.g. decision trees [27] or random forests [28]); and the use of ontologies to find hierarchically related concepts (e.g. patients with closely related diagnosis [29]). However, the temporal dimension of the patient journeys is often not addressed by these algorithms, which requires a broader exploration.

Previous work using temporal information encoded clinical events into streams of information, enabling the use of sequence alignment algorithms in bioinformatics pipelines. Recently, Ledieu et al. [30] used a modified version of the Smith-Waterman sequence alignment algorithm for pharmacovigilance in electronic medical records, detecting inadequate treatment decisions in patient sequences. Meanwhile, Lee & Das [31] developed a temporal sequence alignment strategy to find HIV patients with similar treatment histories, leveraging a simple ontology to improve local alignment of antiretroviral regimens. The use of the ontology certainly adds value to the algorithm, but it is also a limitation if the ontology is not well defined or unknown, for example in the case of the PANS patients.

In this study, we retrospectively investigated the medication prescription patterns of a PANS cohort from Stanford University to elucidate groups of patients with similar treatment patterns. We developed a medication alignment algorithm (*MedAl*) for finding patients with similar patterns, and cluster them to associate their overall impairment scores. Our goal was to facilitate hypothesis generation, for the clinical team and basic science team, that provides guidance on how to better cluster patients for future research and trials.

## 2. Materials and Methods

This is a retrospective cohort study, using patient/parent questionnaires and ledgers collected routinely as part of clinical care, as well as electronic medical records (EMR) data. The objective was to understand patterns of medication usage in patients. Our study involved extracting data from electronic health records, using MedAl to create a patient similarity metric, grouping patients into clusters, visualizing them, evaluating the cluster assignment, and validating the clusters with clinical outcomes in the PANS cohort. The study outline is shown in Figure 1, where our algorithm MedAl allows for downstream clustering by defining a distance between the study subjects.

**Figure 1.**
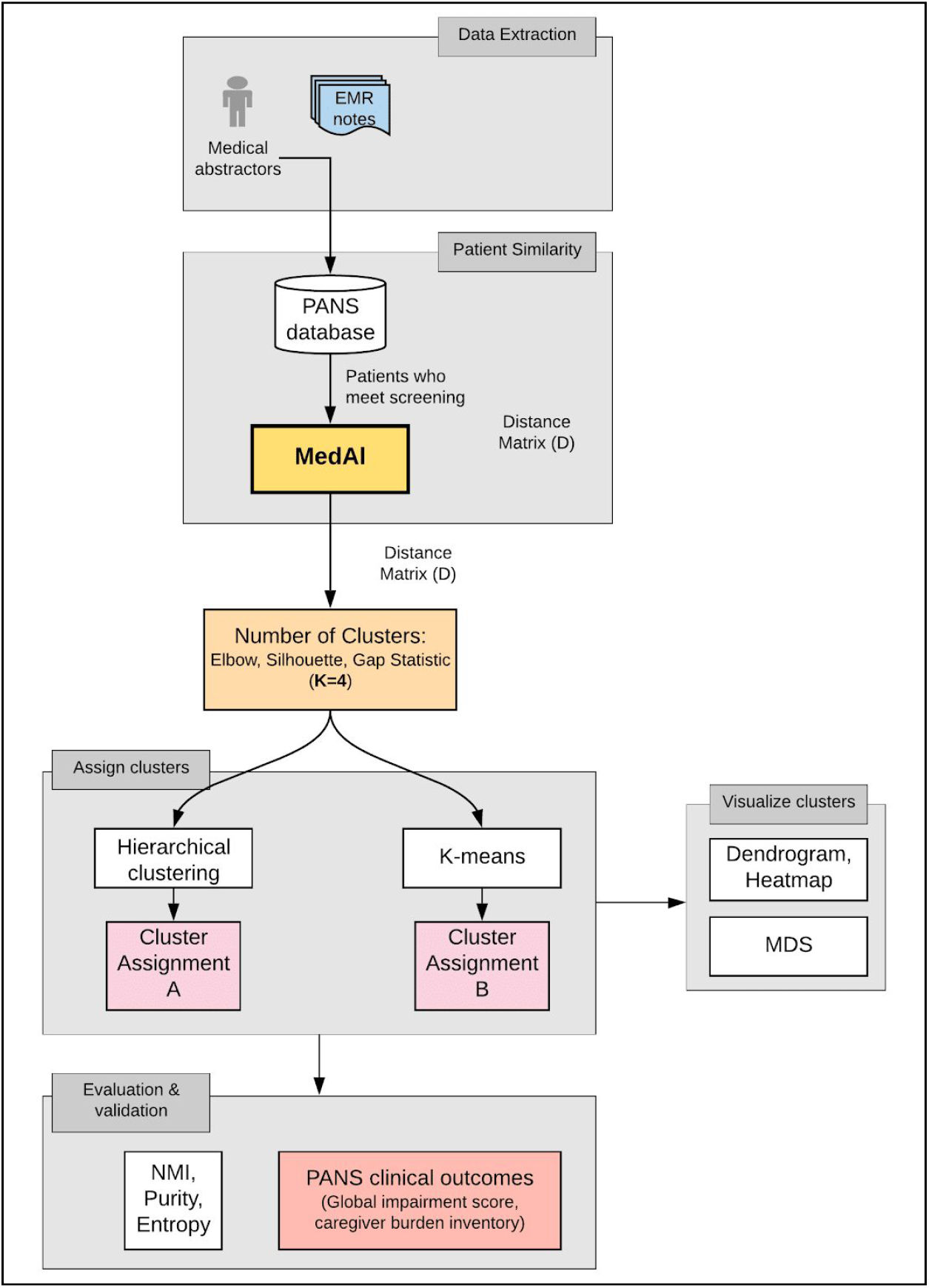
Study outline.

### Clinical setting and population

The pilot study took place at the Stanford PANS clinic located in California, United States of America, which is a multidisciplinary clinic staffed by practitioners of various disciplines (psychiatry, primary care, rheumatology, immunology), as well as a social work psychotherapist and an education specialist.

The data used in this study were generated during clinical care and includes prospectively collected impairment scores (global impairment and caregiver burden inventory), but we also retrospectively collected medication use data abstracted from the clinical charts. Data were collected on those patients seen between the clinic inception on September 3rd, 2012, and the patient cohort identification date for this study, January 31st, 2018. In this period there were 305 patients seen in clinic, out of which we excluded: those whose parents refused consent for the study (N = 4), those patients who did not meet the strict PANS criteria [9] (N = 97), had fewer than three visits to the PANS clinic (N = 36), were older than 12 years at clinic entry (N = 60) and presented to the clinic more than 4 months after onset of psychiatric symptoms (N = 65). The final cohort includes 43 pre-pubertal new-onset patients, as shown in Figure 2. One patient was dropped from the primary analysis because the only medication was taken after the first two years at clinic.

**Figure 2.**
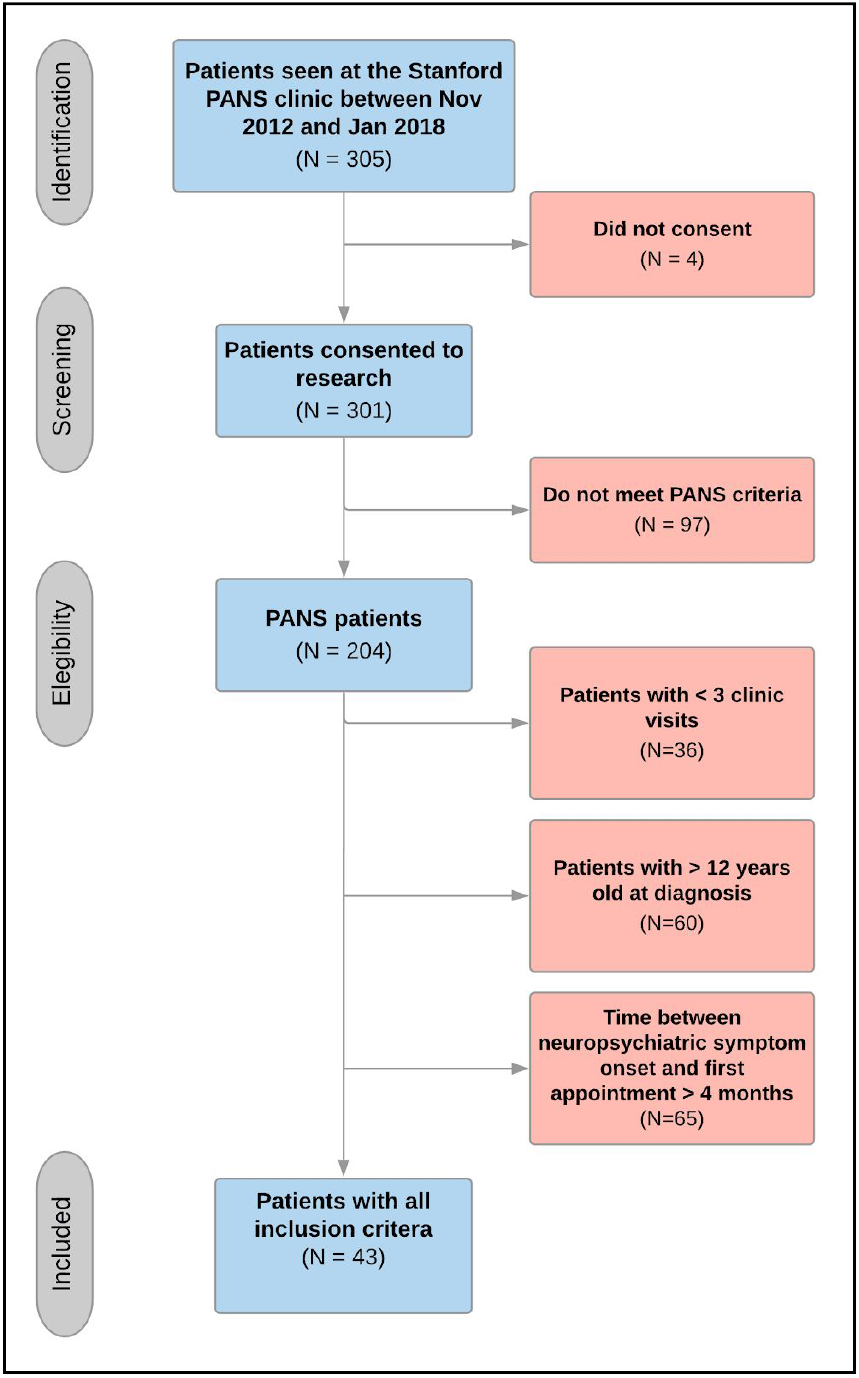
PANS attrition diagram with eligibility criteria.

We only included patients with established care with the clinic (at least three visits) because the first few visits aim to understand the history and disease course, to examine patients and to counsel parents about different treatment options. We used a cut-off age of 12 years to include only pre-pubertal children, as hormones might play a role in psychiatric symptoms and behaviors [32]. We restricted patients to those with such a short time difference between PANS onset and first PANS clinic appointment because we wanted to analyze patients with new-onset PANS, for whom we had more complete information close to the beginning of their PANS illness.

### Data sources

We collected data on medical treatment from the electronic medical record using a keyword search method [33], and we made a final update of medication history in mid-October 2018. For this patient group, a limited number of medications are offered and were decided upon a priori by the clinical team. We excluded short courses (less than 21 days) of antibiotics for acute infections, NSAID taken as needed by patients, and psychiatric medications as the aim of this analysis was to study the similarity of using medical (non-psychiatric/non-psychological) therapies in this group of patients. Our medication keyword search method is outlined in Table 1.

**Table 1.**
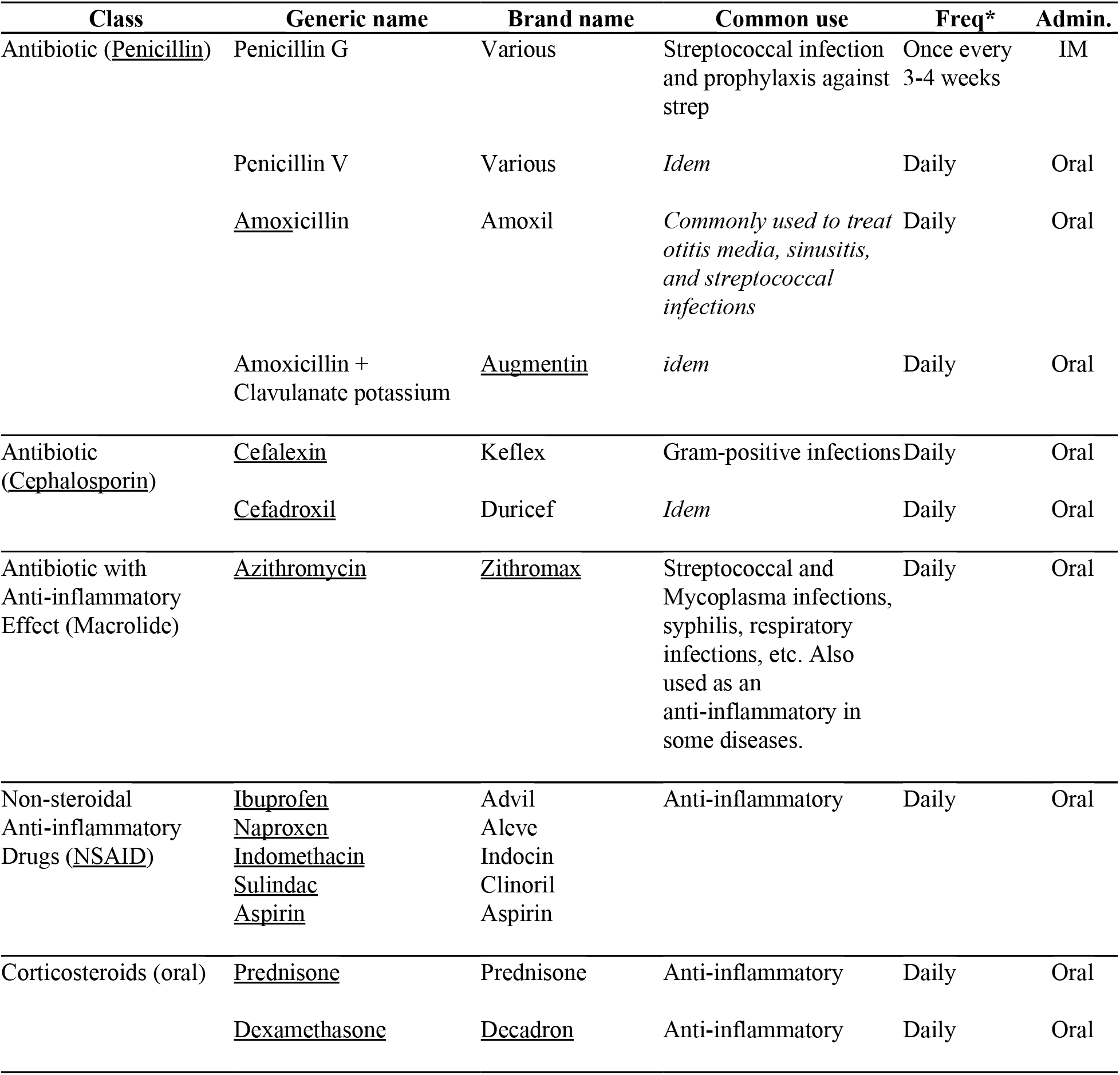
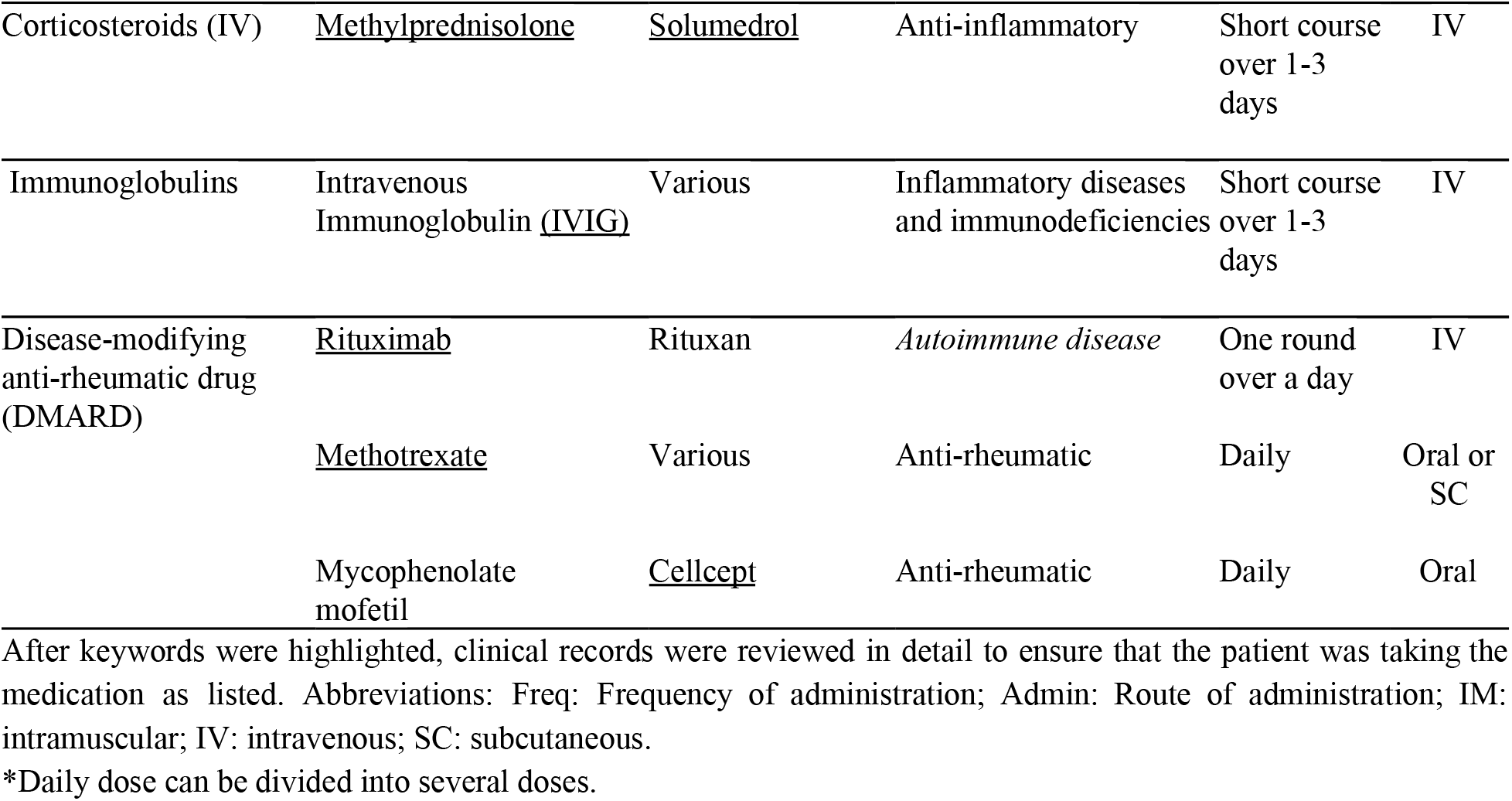
Medication list and keywords (underlined) used in the EMR search-box.

For medications taken daily, we determined the start and stop dates. For medications with long acting effects (i.e. Rituximab, IV corticosteroids) we determined the start date only. In some cases, determining start and stop dates is challenging. For example, a patient/parent may have decided to discontinue a medication between two clinic visits but failed to recall the exact stop date. In these cases, we estimated the stop date using one of two methods: a) if the provider estimated a unit of time during which patient stopped taking the medication, we used the midpoint of that unit of time (e.g. “patient stopped NSAIDs March 2017” would be coded as March 15, 2017; early March will be coded as March 1; late March will be coded as March 30; two weeks ago will be 14 days before the encounter date); b) if no estimate was given, we used the clinic visit date on which the provider reported the patient stopped taking the medication as the stop date. All stop dates for active medications were set at the last visit dates. If a patient suspended the drug for less than a week, we would consider it to be a continuous use; otherwise, we would state two separate periods.

### Patient outcomes

In the PANS cohort, like in many other psychiatric syndromes, evaluating outcomes is a complex task which requires the use of scales and assessment scores that are subjective in nature. The Stanford PANS clinic utilizes two parent rated impairment scales that have been validated in this patient population:

> Global Impairment Scale [34].
>
> A scale ranging from 0 to 100, where the highest value indicates severe challenges for the patient to interact with others and carry on their daily activities, and the lowest value indicates a child without impairment.
>
> Caregiver Burden Inventory [35].
>
> A scale ranging from 0 to 96, where the highest value indicates the greatest caregiver burden. A score greater than 36 indicates respite care [36].

The Stanford PANS clinic collects electronic patient questionnaires that caregivers and patients fill out before each clinic visit. The questionnaire queries symptom-specific scales corresponding to the severity of the patient symptoms. The outcome data (Global Impairment and Caregiver Burden) may be skewed by the fact that the frequency of follow-up clinic visits and corresponding completed questionnaires trends with the severity of the psychiatric symptoms (i.e. when patients are highly symptomatic, they come to the clinic weekly for psychiatric medication titration and therapy, but when the child improves/resolves their psychiatric symptoms, it is difficult to get the family to return to clinic).

### A modified sequence alignment algorithm

In this manuscript, we propose medication alignment for patient similarity (*MedAl*) algorithm, that adapts a protein sequence alignment paradigm [37, 38] to medication usage history. The alignment edit distance is used to estimate medication usage similarity, in order to construct a clustering strategy. The algorithm is as follows: a) encode the medication history in a sequence representation; b) perform alignment of pairs of medication sequences; and c) compute a weighted patient pairwise edit distance.

#### Encode the medication history

For every pair of patients taking the same medication, usage was encoded into a binary vector representing daily intake. Thus, the dimension of the vector is determined by the earliest and latest day of medication usage among both patients. We considered missing values to be not missing at random (NMAR), and therefore assigned an explicit missing value.

#### Sequence alignment

We used a dynamic programming approach to align the vectors generated in the previous step. First, an alignment matrix is generated, with the sequence for the one patient as column names, and the sequence for the other patient as row names. The matrix is initialized with zeros in the first column and row. Then, the matrix is filled with values considering at each cell only the three neighbors of that cell (left, upper-left diagonal, and top cell), starting from the top-left of the matrix, to the bottom-right.

MedAl considers four medication usage patterns to fill values at each cell: start of medication, continuing medication, end of medication, and continuing gap. For any pair of patients, there are 16 possible combinations for how the medication is being administered. Four rules are used to assign values in each cell of the matrix: 1) if both patients have the same medication usage pattern, then the minimum neighboring value is assigned; 2) if one of the patients has switching pattern (e.g. the first patient starting medication, and second patient continuing medication), then the maximum neighboring value is assigned; 3) if both patients have complete opposing patterns (e.g. first patient is starting medication while the second patient is ending medication), then the maximum neighboring value plus one is assigned; and 4) lastly, if one of the patients has a pattern involving missing values (e.g. first patient is starting medication, while the second patient is not taking anything), then the minimum neighboring value plus one is assigned.

#### Weighted edit distance

The edit distance is a metric of the minimum number of operations needed to align two sequences. Traditionally in genome sequences, these operations could be insertion of a gap, deletion of a position, or substitution for an equivalent letter. However, for medication alignment, we only allow insertions, since we do not want to delete medication histories, and a substitution for an equivalent letter is difficult to assess. In Figure 3, the minimum distance, for a toy example, is shown as the green path.

**Figure 3.**
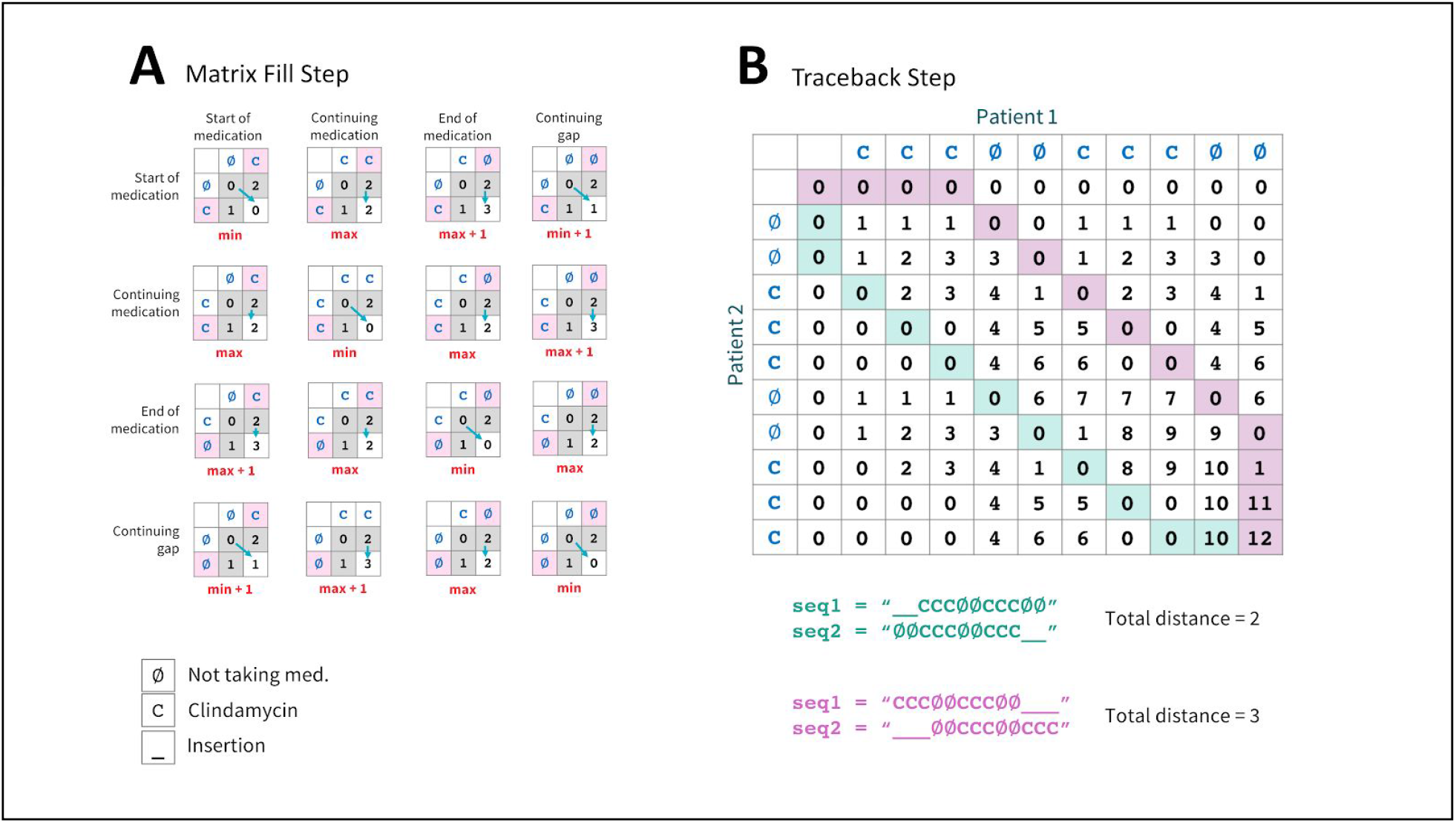
Dynamic programming approach to medication alignment. A) matrix fill step; B) Traceback step. The path starts from the bottom-right cell and ends at the top-left cell of the matrix. Only two insertions were needed to align the sequences. The purple path is an alternative sub-optimal alignment that required three insertions, but this distance is not selected.

### Cluster assignment and evaluation

Two methods were used for clustering: hierarchical clustering and k-means.

> *Hierarchical clustering*. This is a popular technique that creates clusters with an ordering (hierarchy). We selected an agglomerative (bottom-up) algorithm with complete linkage. This method starts by assigning each sample to its own cluster. Then, to join the two most similar patients into a larger cluster, we used the *MedAl* distance. The algorithm continues to recursively aggregate clusters until all samples have been added to a single cluster.
>
> *K-means*. This clustering method is also a popular clustering method which has been previously used in psychiatric disorders [39]. This technique aims to partition observations of patients into *k* clusters in which each observation belongs to the cluster with the nearest mean. The metric to estimate distance is typically a geometrical distance (e.g. Euclidean distance). This metric was not feasible with the medication history data, therefore *MedAl* was used.

For comparison between these two clustering methods, we used three metrics using the normalized mutual information (NMI), cluster purity, and cluster entropy methods. All three methods range from 0, being the worst evaluation, to 1, being the best evaluation. The selection of k=6 seems to be the best balance across all three metrics. For the PANS cohort, the ground truth of cluster assignment is unknown, and therefore these values only serve to provide confidence on the equivalence of clustering assignment between both methods..

Four quantitative methods were used to estimate the optimal number of clusters (*k*) in the PANS cohort: a) elbow method, which measures the compactness of clusters by summing the within-cluster sum of squares; b) Silhouette method [40], which measures the cohesion of clusters, through averaging the distance between each element and the rest of the elements in that cluster, and comparing to the average distance of neighboring clusters; c) Gap statistic [41], which estimates the statistical comparison between the total intra-cluster variation and the null hypothesis without cluster assignment; and d) Clest method [42], which is a variation on the gap statistic, selecting the number of clusters within a range from the maximum global standard error.

To compare trends between clusters for the Global Impairment score and Caregiver Burden Inventory, we conducted an analysis of variance (ANOVA) accounting for longitudinal sampling from each individual. Slopes were fit stratified by cluster using a linear mixed-effects model to determine trends within clusters for both scores.

## 3. Results

### Clinical characteristics

Patient characteristics in this study were evenly distributed by sex, and heavily skewed by self-reported race and ethnicity (mostly non-Hispanic white) in patients. The age at first neuropsychiatric deterioration occurred on average between 7-8 years old, with a rapid intake by the PANS clinic (patients seen > 4 months after psychiatric symptom onset were excluded). Table 2 shows a detailed description of the clinical characteristics of this cohort.

**Table 2.** Clinical characteristics of 43 consecutive pre-pubertal patients with new-onset PANS.

| Characteristic | PANS |
| --- | --- |
| **Overall** | **N = 43** |
| **Sex** |  |
| Female | 23 (53.49%) |
| Male | 20 (46.51%) |
| **Age at first neuropsychiatric symptom onset (years)** | mean = 7.77<br>SD = 2.35 |
| **Age at first clinic visit (years)** | mean = 7.93<br>SD = 2.39 |
| **Race / ethnicity\*** |  |
| Non-Hispanic White | 39 (90.7%) |
| Non-Hispanic Asian | 4 (9.3%) |
| Hispanic / Latino | 4 (9.3%) |
| Other / unknown | 4 (4.65%) |
| **PANS symptoms at first presentation to the clinic** |  |
| Obsessive compulsive symptoms | 36 (83.72%) |
| Food intake problems | 18 (41.86%) |
| Anxiety / phobia | 37 (86.05%) |
| Emotional lability / depression / suicidal ideation | 33 (76.74%) |
| Aggression / irritability / opposition | 33 (76.74%) |
| Cognitive problems | 9 (20.93%) |
| Inattention / deterioration in school | 24 (55.81%) |
| Behavioral regression | 27 (62.79%) |
| Sensory amplification | 19 (44.19%) |
| Sleep disturbances | 26 (60.47%) |
| Urinary problems | 14 (32.56%) |
| Motor / vocal tics | 24 (55.81%) |

### Number of Clusters

A graphical inspection of the methods in Figure 4 shows disagreement between them. The elbow method does not clearly show an elbow, suggesting that any number of clusters would show some effect. In the Silhouette method, the maximum value was at k=9 (the highest rated), however, there are two good candidates at k=2 and k=4. The gap statistic method shows the best value at k=2. The Clest method suggests a number of clusters of k=6. On the other hand, when comparing cluster purity, NMI and Entropy, these metrics seem to converge around k=6.

**Figure 4.**
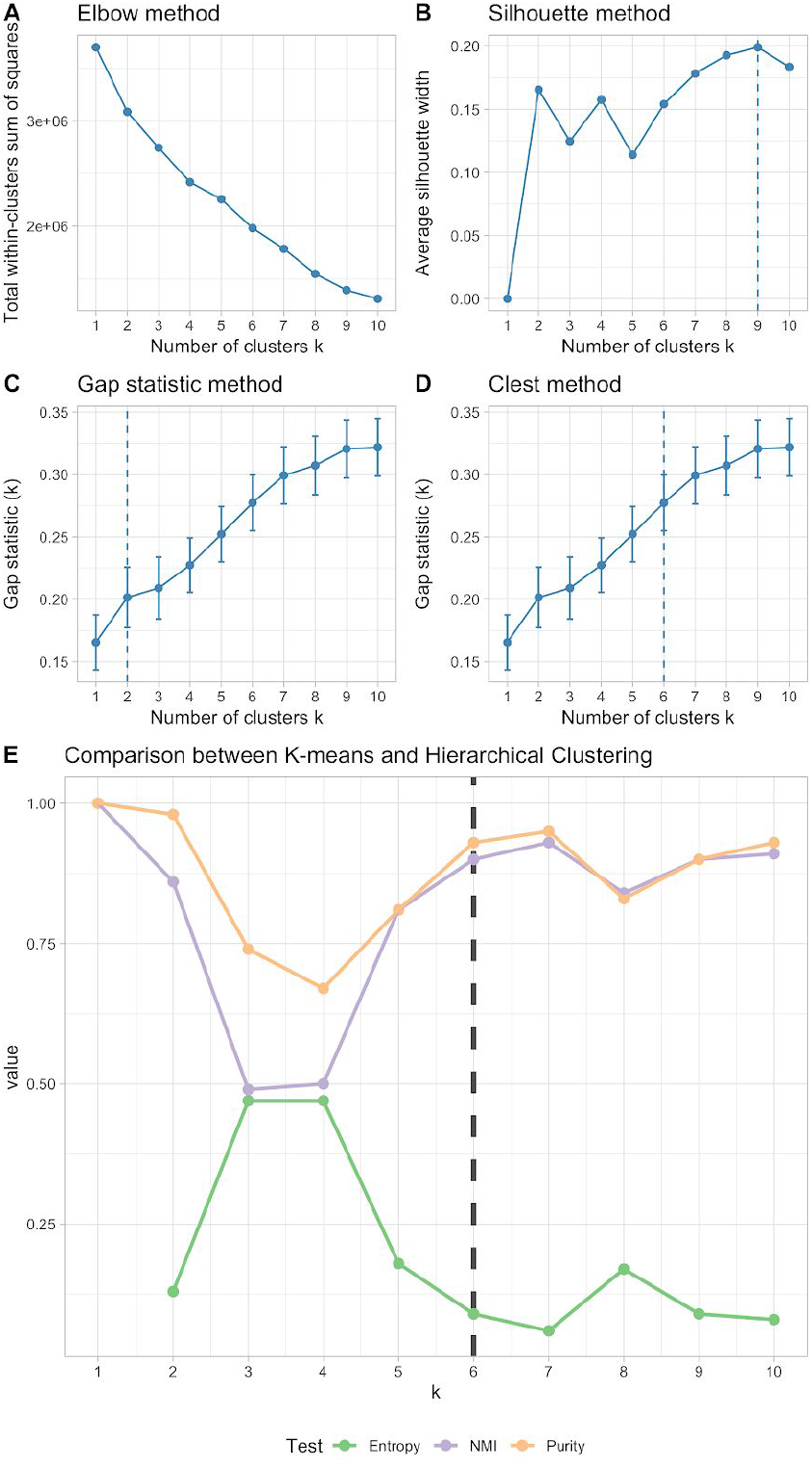
Analysis of optimal number of clusters. A) Elbow method, B) Silhouette method, C) Gap statistic method, D) Clest method, E) Evaluation of normalized mutual information (NMI), Purity and Entropy.

### Cluster assignment

With no clear selection of the best number of clusters, but with some good candidates (k=2, 4, 6, 9), we used another visualization tool, showing the MedAl similarity metric in a heatmap with the corresponding dendrogram and annotations of both the k-means and hierarchical clustering (hclust) methods, as shown in Figure 5. For the remainder of the analysis we used k=6, given that it was the number of clusters that most clearly differentiated between the groups.

**Figure 5.**
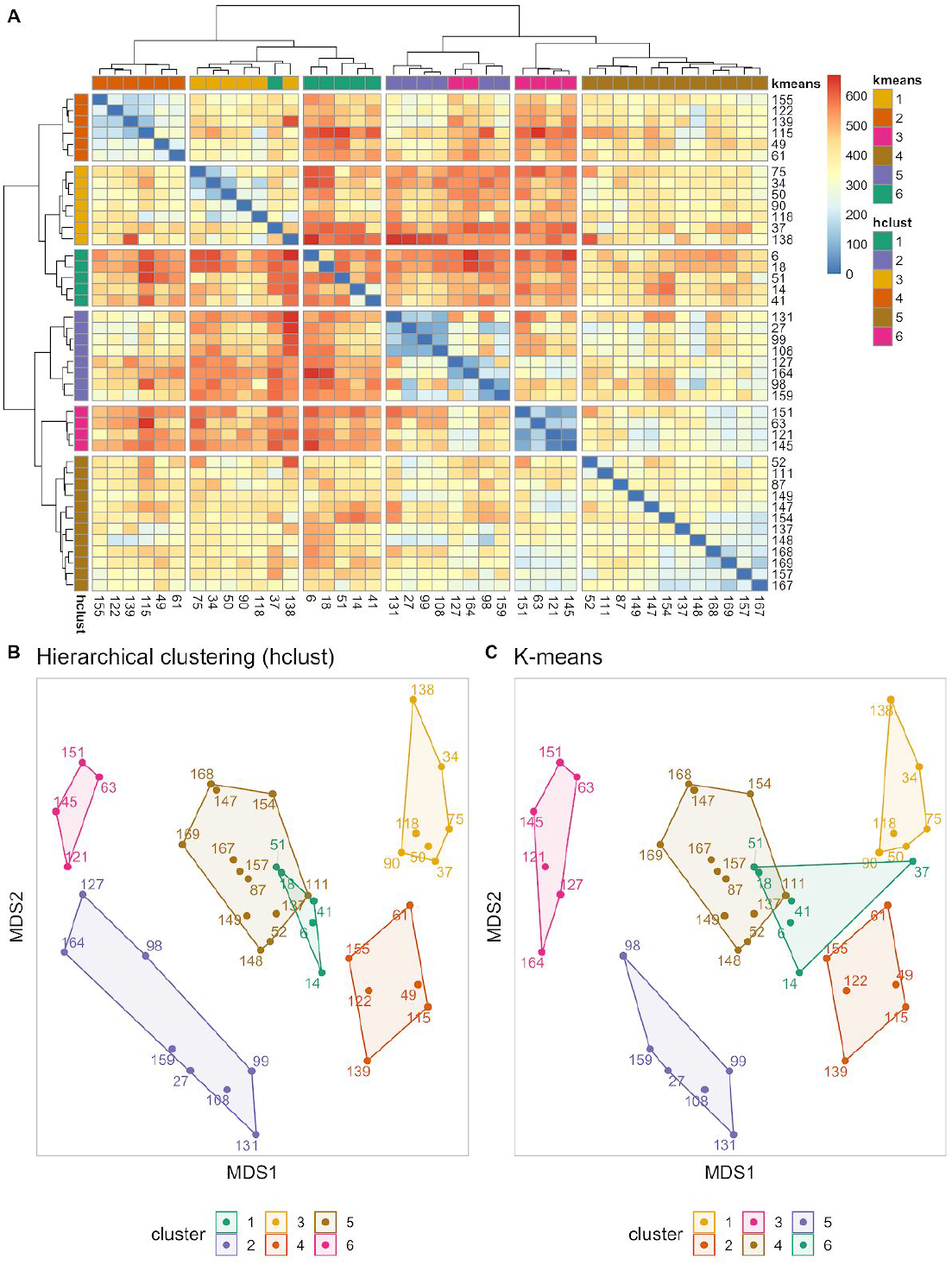
Cluster assignment for hierarchical clustering using k = 6. A) Heatmap of the MedAl score, clustering dendrogram in rows (hierarchical clustering, hclust) and columns (k-means); B) MDS components 1 and 2 with hierarchical clustering assignment; C) MDS components 1 and 2 with k-means assignment.

### Cluster characteristics

We also assessed the qualitative value of the clustering assignment, based on the patient similarity metric calculated using *MedAl*. The medication usage by cluster and medication class is shown in Figure 6. The patients were censored at two years of follow-up, and the index date used to align all medication histories is the first visit to the PANS clinic. Each medication class varies from none of the patients taking that drug to all of the patients taking the drug. Clusters 4 and 6, composed by patients with the most severe groups requiring high dose IV steroids; Clusters 1 and 2, composed by patients requiring prolonged NSAID which are used frequently by arthritic or inflammatory diseases; Cluster 2 was also a group of patients with likely underlying Streptococcal infections treated with penicillin; Clusters 3 and 5, composed by patients taking a short course of NSAID for their relapsing/remitting courses of PANS illness. The psychometric scores as outcomes in each cluster generally improved within the first two years except the group of patients using prolonged NSAID and likely suffering from arthritic or inflammatory diseases that ultimately required more aggressive immunomodulation.

**Figure 6.**
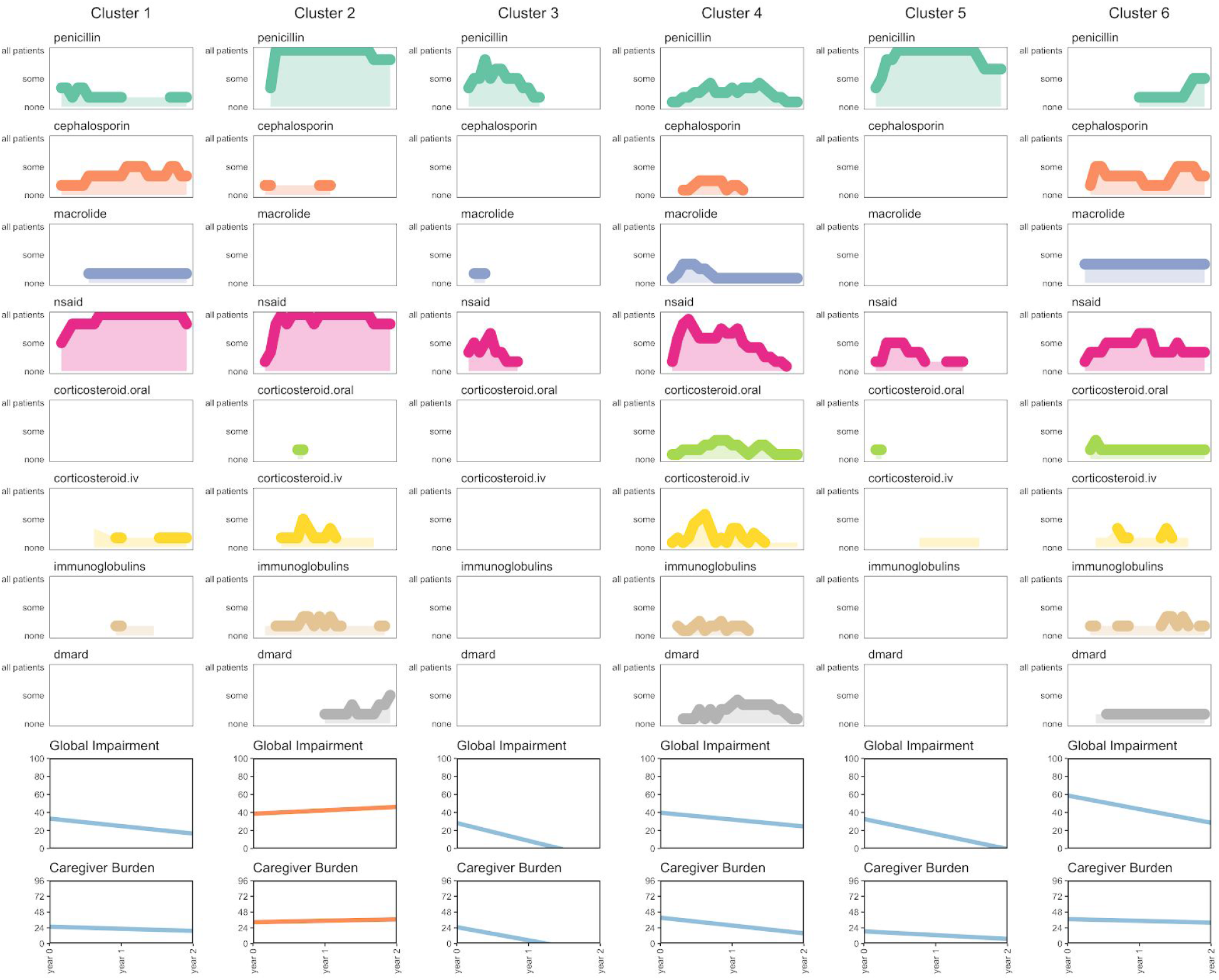
Drug usage of patients in clusters and overall impairment scales. Abbreviations: dmard: disease-modifying antirheumatic drug; nsaid: Nonsteroidal anti-inflammatory drug. For a full list of drug categories, please refer to Table 1. The last two rows show the Global Impairment Scale and Caregiver Burden Inventory, where linear model fitting for all patients in that cluster, and the color corresponds to a positive slope (red) meaning worsening of the patient; and a negative slope (blue), meaning an improvement on the patient’s impairment or caregiver burden. The cluster assignment was obtained from k-means.

Upon performing an analysis of variance (ANOVA), we found in Table 3 that there is significant heterogeneity in trend of scores over time within an individual and between clusters for both Global Impairment (P_ANOVA_=0.005) and Caregiver Burden (P_ANOVA_=0.0004). We note that all clusters, with the exception of cluster 2, demonstrate on average a decreased Caregiver Burden Inventory and decreased Global Impairment Scale over time. This is a sign of general improvement on the patient’s condition.

**Table 3.** Slopes for Caregiver Burden Inventory and Global Impairment scores over time by cluster.

|  | Global Impairment Score |  |  |  | Caregiver Burden |  |  |  |
| --- | --- | --- | --- | --- | --- | --- | --- | --- |
|  | *Intercept* | *Slope* | *Std. Error* | *P-value* | *Intercept* | *Slope* | *Std. Error* | *P-value* |
| **Cluster 1** | 33.20 | -8.32 | 5.88 | 0.2234 | 25.78 | -3.20 | 4.17 | 0.4595 |
| **Cluster 2** | 38.58 | 3.80 | 2.65 | 0.1562 | 32.60 | 2.17 | 5.05 | 0.6845 |
| **Cluster 3** | 28.24 | -19.75 | 4.31 | 0.0002 | 25.05 | -20.10 | 5.55 | 0.0029 |
| **Cluster 4** | 40.46 | -8.89 | 5.13 | 0.1140 | 39.42 | -11.90 | 3.13 | 0.0005 |
| **Cluster 5** | 32.72 | -16.67 | 8.52 | 0.1060 | 18.69 | -5.80 | 3.56 | 0.1135 |
| **Cluster 6** | 58.80 | -15.14 | 3.25 | 0.0006 | 37.33 | -2.70 | 1.94 | 0.1734 |
| ANOVA P-value = **0.0054** |  |  |  |  | ANOVA P-value = **0.0004** |  |  |  |

## 4. Discussion

Here we present a method to align patient history to identify distinct clusters of medication usage, and we apply the method to a PANS cohort. Our approach relies on constructing a symmetric matrix of the pairwise distances between patients. The dynamic programming approach used for constructing the distance between each pair scales as the square of the time period under consideration. Therefore, construction of the distance matrix for *N* patients taking *D* drugs over a period of length *T* takes *O*(*DN^2^T^2^*) time and uses *O*(*N^2^* + *T^2^*) memory. In cases where the exact intervals and number of days that the drug is administered is not crucial the time periods can be collapsed (e.g. from days to weeks) in order to increase computational and memory efficiency.

Our strategy identifies clusters to characterize the PANS patient population, but cluster assignment should not be considered an association without further investigation. This approach is merely a way to generate new hypotheses that could be further investigated by the clinical and research teams. In a polypharmacy context, this approach could further be used to better understand treatment patterns in the clinic. These patterns may represent clinician’s preference of treatments for certain clinical characteristics and severity levels, or effectiveness of a treatment such that it is continuously prescribed.

We identified six apparent clusters in the PANS cohort based on their medication history, including initiation and cessation dates. These clusters can be further grouped into three groups: a) those patients requiring high dose IV steroids (clusters 4 and 6); b) patients with higher arthritic or inflammatory burden requiring prolonged NSAID (clusters 1 and 2), where one was also treated with penicillins presumably due to concerns for Streptococcal infections (cluster 2); and c) patients with mild relapsing/remitting syndrome treated with a short course of NSAID (clusters 3 and 5).

The peak of IV corticosteroid use in Clusters 2, 4 and 6, was followed by a steady increase in use of several other medications (steroid sparing agents including DMARDS and IVIG). This pattern is consistent with a more severe phenotype since the clinicians in the PANS clinic reserve IV corticosteroids for the most severe cases, who tend to also have comorbid arthritis and other autoimmune diseases. This is supported by the outcomes data (Figure 6, last two rows). At clinic entry, the patients in these clusters had a starting Global Impairment score above 40 (in a scale up to 100). However, the outcomes for these clusters were very different. Cluster 2 generally worsened throughout the two years of study except perhaps after the peak use of IV corticosteroids; meanwhile Clusters 4 and 6 generally improved overall. This differentiation on outcomes shows that our clustering strategy accurately captured the decision making process by the clinician when prescribing other drugs to account for the patient’s health status at the time of prescription.

Constant heavy use of NSAIDs in Clusters 1, 2 may represent a subgroup of patients who respond to NSAIDs but rely on the constant use of NSAIDS to suppress symptoms. This continued long term usage may reflect those patients who had recrudescence of symptoms when the NSAID dose was lowered. However, this group seemed to worsen over time which appeared to be associated with the eventual addition of more aggressive immunomodulation. The low Global Impairment score at clinic entry of Cluster 1 may have led to the decision not to use corticosteroids in the initial treatment. In contrast, short courses of NSAID in Clusters 3 and 5, significantly improved the psychometric scores significantly during the first half year. This pattern is compatible with the relapsing/remitting course of PANS illness. When the patients returned to near baseline, clinicians stopped most pharmacological treatments.

Heavy penicillin use was observed in Clusters 2 and 5 (also moderate use in Cluster 3) which may indicate that the clinician had suspicion or evidence for a streptococcal infection, coincident with the onset of the psychiatric illness (an association seen in epidemiological studies [9]) and likely reflects the clinicians attempt to use penicillin as prophylaxis against streptococcus. Cluster 2 had the highest penicillin usage of all clusters, and given the increase in the psychometric score burden likely reflects relapse of these patients, and appears to be coincident with decreasing penicillin usage.

Our study was limited by a small number of patients selected for this study. The selection of patients was reduced from an initial cohort of 305 patients seen in the Stanford PANS clinic to 43 pre-pubescent patients who met strict PANS criteria and who were new-onset at the time of clinic entry, such that their medication history was fully revealed in the charts to avoid recall bias and unclear history. Furthermore, we limited the timespan of medication history to only the first two years of treatment, to make the patient comparisons possible. Additionally, we grouped medications of similar effects into clinically relevant categories instead of treating each drug independently to increase power and applicability to the clinical practice. The stricter study criteria and limited time frame increased comparability between patients to find patterns related to medication usage immediately after diagnosis and treatment initiation. In future research, selecting an appropriate censoring strategy and time zero should be individualized to research questions, as complexity of patients’ medical and drug history often exist in real-world data studies.

The field of patient similarity is expanding with the inclusion of novel sources of data in electronic format. In this study, we have shown that *MedAl* is capable of providing a reliable similarity metric that can generate new hypotheses for further investigation in a complex syndrome like PANS. The medication histories of other cohorts with a high pharmacological burden will play an important role in our understanding of their treatment patterns.

## Data Availability

The data that supports the findings of this study is available for research purposes upon written request, which will be reviewed on a case-by-case basis by Dr. Jennifer Frankovich, director of the Stanford PANS clinic (http://med.stanford.edu/pans), and Stanford's Institutional Review Board (IRB).

http://med.stanford.edu/pans.html

## Contributions

ALP designed the study. JF designed the patient inclusion/exclusion criteria, designed the medication categories and medication search strategy, designed the prospective clinical questionnaires and outcome scores and supervised the clinical database and medication abstraction and entry. CMCL, AC, and JF reviewed medical notes and drafted part of the manuscript. GLW, AP, AI, and ALP performed analysis of data. CDB, GLW, and JF provided interpretation of the results. ALP drafted the manuscript, and all authors contributed critically, read, revised and approved the final version.

## Ethics approval

This study was approved by Stanford University’s Institutional Review Board (IRB). Written informed consents were obtained from parents of minors, and assents were obtained from subjects aged 7 to 17 (all patients in this cohort) who were able to understand it (IRB protocol #26922). We did not have the opportunity to re-consent two patients at the time of data abstraction and analysis. The inclusion of their data is covered by a retrospective chart review protocol (IRB protocol #28533). Authors CMCL, AC, and JF served as honest brokers securing identifiable information. The remaining authors only had access to de-identified information. Non-human subject determination was provided (IRB protocol #46979).

## Code Availability

The MedAl and PyMedAl implementation can be accessed through the public repository: https://github.com/bustamante-lab/medal

## References

1. Masnoon, N., Shakib, S., Kalisch-Ellett, L. & Caughey, G. E. What is polypharmacy? A systematic review of definitions. BMC Geriatr 17, 230 (2017).

2. Shah, B. M., & Hajjar, E. R. Polypharmacy, adverse drug reactions, and geriatric syndromes. Clin. Geriatr. Med. 28, 173–186 (2012).

3. Morandi, A. et al. Predictors of rehospitalization among elderly patients admitted to a rehabilitation hospital: the role of polypharmacy, functional status, and length of stay. J Am Med Dir Assoc 14, 761–767 (2013).

4. Díaz-Caneja, C. M., Espliego, A., Parellada, M., Arango, C. & Moreno, C. Polypharmacy with antidepressants in children and adolescents. Int. J. Neuropsychopharmacol. 17, 1063–1082 (2014).

5. Marengo, M. F. et al. Measuring therapeutic adherence in systemic lupus erythematosus with electronic monitoring. Lupus 21, 1158–1165 (2012).

6. Justice, A. C. et al. Nonantiretroviral polypharmacy and adverse health outcomes among HIV-infected and uninfected individuals. AIDS 32, 739–749 (2018).

7. Alshamrani, M., Almalki, A., Qureshi, M., Yusuf, O. & Ismail, S. Polypharmacy and Medication-Related Problems in Hemodialysis Patients: A Call for Deprescribing. Pharmacy (Basel) 6, (2018).

8. LeBlanc, T. W., McNeil, M. J., Kamal, A. H., Currow, D. C. & Abernethy, A. P. Polypharmacy in patients with advanced cancer and the role of medication discontinuation. Lancet Oncol. 16, e333–41 (2015).

9. Swedo, S. E. et al. Clinical presentation of pediatric autoimmune neuropsychiatric disorders associated with streptococcal infections in research and community settings. J Child Adolesc Psychopharmacol 25, 26–30 (2015).

10. Brown, K. et al. Pediatric Acute-Onset Neuropsychiatric Syndrome Response to Oral Corticosteroid Bursts: An Observational Study of Patients in an Academic Community-Based PANS Clinic. J Child Adolesc Psychopharmacol 27, 629–639 (2017).

11. Zheng J, Frankovich J, McKenna E, Rowe N, MacEachern S, Ng N, et al. Association of Pediatric Acute-Onset Neuropsychiatric Syndrome With Microstructural Differences in Brain Regions Detected via Diffusion-Weighted Magnetic Resonance Imaging. JAMA Network Open 3(5) (2020)

12. Giedd JN, Rapoport JL, Garvey MA, Perlmutter S, Swedo SE. MRI assessment of children with obsessive-compulsive disorder or tics associated with streptococcal infection. Am J Psychiatry J Psychiatry. 157(2), 281–3 (2000)

13. Kumar A, Williams MT, Chugani HT. Evaluation of basal ganglia and thalamic inflammation in children with pediatric autoimmune neuropsychiatric disorders associated with streptococcal infection and tourette syndrome: a positron emission tomographic (PET) study using 11C-[R]-PK11195. J Child Neurol. 30(6), 749–56 (2015).

14. Lotan, D., Benhar, I., Alvarez, K., Mascaro-Blanco, A., Brimberg, L., Frenkel, D., Cunningham, M.W. and Joel, D. Behavioral and neural effects of intra-striatal infusion of anti-streptococcal antibodies in rats. Brain, behavior, and immunity, 38, 249–262.(2014)

15. Brimberg L, Benhar I, Mascaro-Blanco A, Alvarez K, Lotan D, Winter C, et al. Behavioral, pharmacological, and immunological abnormalities after streptococcal exposure: A novel rat model of sydenham chorea and related neuropsychiatric disorders. Neuropsychopharmacology. 37(9), 2076–87 (2012).

16. Frankovich, J. et al. Multidisciplinary clinic dedicated to treating youth with pediatric acute-onset neuropsychiatric syndrome: presenting characteristics of the first 47 consecutive patients. J Child Adolesc Psychopharmacol 25, 38–47 (2015).

17. Swedo, S. E., Frankovich, J. & Murphy, T. K. Overview of Treatment of Pediatric Acute-Onset Neuropsychiatric Syndrome. J Child Adolesc Psychopharmacol 27, 562–565 (2017).

18. Brown, S.-A. Patient Similarity: Emerging Concepts in Systems and Precision Medicine. Front. Physiol. 7, 19 (2016).

19. Sharafoddini, A., Dubin, J. A. & Lee, J. Patient Similarity in Prediction Models Based on Health Data: A Scoping Review. JMIR Med Inform 5, e7 (2017).

20. Cahan, A. & Cimino, J. J. Visual assessment of the similarity between a patient and trial population: Is This Clinical Trial Applicable to My Patient? Appl Clin Inform 7, 477–488 (2016).

21. Li, L. et al. Identification of type 2 diabetes subgroups through topological analysis of patient similarity. Sci Transl Med 7, 311ra174–311ra174 (2015).

22. Panahiazar, M., Taslimitehrani, V., Pereira, N. L. & Pathak, J. Using EHRs for Heart Failure Therapy Recommendation Using Multidimensional Patient Similarity Analytics. Stud Health Technol Inform 210, 369–373 (2015).

23. Lee, J. et al. Privacy-Preserving Patient Similarity Learning in a Federated Environment: Development and Analysis. JMIR Med Inform 6, e20 (2018).

24. Zhang, P., Wang, F., Hu, J. & Sorrentino, R. Towards personalized medicine: leveraging patient similarity and drug similarity analytics. AMIA Jt Summits Transl Sci Proc 2014, 132–136 (2014).

25. Dai, Y., Lokhandwala, S., Long, W., Mark, R. & Lehman, L.-W. H. Phenotyping Hypotensive Patients in Critical Care Using Hospital Discharge Summaries. IEEE EMBS Int Conf Biomed Health Inform 2017, 401–404 (2017).

26. Ng, K., Sun, J., Hu, J. & Wang, F. Personalized Predictive Modeling and Risk Factor Identification using Patient Similarity. AMIA Jt Summits Transl Sci Proc 2015, 132–136 (2015).

27. Wang, F. Adaptive semi-supervised recursive tree partitioning: The ART towards large scale patient indexing in personalized healthcare. Journal of Biomedical Informatics 55, 41–54 (2015).

28. Lee, J. Patient-Specific Predictive Modeling Using Random Forests: An Observational Study for the Critically Ill. JMIR Med Inform 5, e3 (2017).

29. Girardi, D. et al. Using concept hierarchies to improve calculation of patient similarity. Journal of Biomedical Informatics 63, 66–73 (2016).

30. Ledieu, T., Bouzille, G., Plaisant, C., Thiessard, F., Polard, E., & Cuggia, M. (2018). Mining clinical big data for drug safety: Detecting inadequate treatment with a DNA sequence alignment algorithm. AMIA Annual Symposium Proceedings, 1368 (2018).

31. Lee, W. N., & Das, A. K. Local alignment tool for clinical history: temporal semantic search of clinical databases. AMIA Annual Symposium Proceedings, 437 (2010).

32. Mitra, S., Bastos, C. P., Bates, K., Pereira, G. S. & Bult-Ito, A. Ovarian Sex Hormones Modulate Compulsive, Affective and Cognitive Functions in A Non-Induced Mouse Model of Obsessive-Compulsive Disorder. Front Behav Neurosci 10, 215 (2016).

33. Frankovich, J., Longhurst, C. A. & Sutherland, S. M. Evidence-based medicine in the EMR era. N Engl J Med 365, 1758–1759 (2011).

34. Leibold, C., Thienemann, M., Farhadian, B., Willett, T. & Frankovich, J. Psychometric Properties of the Pediatric Acute-Onset Neuropsychiatric Syndrome Global Impairment Score in Children and Adolescents with Pediatric Acute-Onset Neuropsychiatric Syndrome. J Child Adolesc Psychopharmacol 29, 41–49 (2019).

35. Farmer, C. et al. Psychometric Evaluation of the Caregiver Burden Inventory in Children and Adolescents With PANS. J Pediatr Psychol 43, 749–757 (2018).

36. Frankovich, J., Leibold, C. M., Farmer, C., Sainani, K., Kamalani, G., Farhadian, B.,... & Thienemann, M. The Burden of Caring for a Child or Adolescent With Pediatric Acute-Onset Neuropsychiatric Syndrome (PANS): An Observational Longitudinal Study. The Journal of clinical psychiatry, 80–1 (2018).

37. Lipman, D. J., & Pearson, W. R. Rapid and sensitive protein similarity searches. Science 227, 1435–1441 (1985).

38. Pearson, W. R. & Lipman, D. J. Improved tools for biological sequence comparison. PNAS 85, 2444–2448 (1988).

39. Fuente-Tomas, L., Arranz, B., Safont, G., Sierra, P., Sanchez-Autet, M., Garcia-Blanco, A., & Garcia-Portilla, M. P. Classification of patients with bipolar disorder using k-means clustering. PloS one, 14–1 (2019).

40. Rousseeuw, P. J. 1987. Silhouettes: A graphical aid to the interpretation and validation of cluster analysis. Journal of computational and applied mathematics 53–65 (1987).

41. Tibshirani, R., Walther, G. & Hastie, T. Estimating the number of clusters in a data set via the gap statistic. Journal of the Royal Statistical Society: Series B (Statistical Methodology) 63, 411–423 (2001).

42. Dudoit, S., & Fridlyand, J. A prediction-based resampling method for estimating the number of clusters in a dataset. Genome biology, 3–7 (2002)

